# SmART-TBI: A fully remote protocol for a randomized placebo-controlled double-blinded clinical trial for a dietary supplement to improve sleep in Veterans

**DOI:** 10.1101/2025.02.22.25322722

**Authors:** Jonathan E. Elliott, Savanah J. Sicard, Cosette Olivo, Hannah A. Cunningham, Arlynn E. Ekis, Katherine L. Powers, Jessica S. Brewer, Jennifer D’Silva, Sarah Happ, Andrea Hildebrand, Akiva Cohen, Miranda M. Lim

## Abstract

Traumatic brain injury (TBI) is associated with chronic sleep disturbances and cognitive impairment, with limited effective therapeutic strategies. Our previous work showed dietary supplementation with branched chain amino acids (BCAAs; isoleucine, leucine, valine), the primary substrate for *de novo* glutamate/GABA synthesis in the CNS, restored normal sleep-wake patterns and improved cognitive function in rodents. Our recent pilot work in humans showed preliminary feasibility/acceptability and limited efficacy for BCAAs to improve sleep in Veterans with TBI. However, these pilot data were limited in sample size, treatment dosages/duration, and therefore unable to establish efficacy or provide insight into dosing/duration parameters. The present study, SmART-TBI (supplementation with amino acid rehabilitative therapy in TBI: NCT04603443), represents a fully powered, placebo-controlled, double-masked randomized clinical trial (target n=120). Covariate adaptive randomization controlling for age, sex, TBI recency, pain, depression, and PTSD, allocated participants 1:1:1:1 to four groups comprising 3 BCAA doses (‘high’ 30g b.i.d.; ‘medium’ 20g b.i.d.; and ‘low’ 10g b.i.d.) and one placebo-control (rice protein, 10g b.i.d.). Outcome measures were assessed following a 2-week baseline period; after 4 weeks, 8 weeks, and 12 weeks of intervention; and after 4 weeks and 12 weeks post-intervention. Primary outcomes were feasibility and acceptability of the protocol. Exploratory outcomes included preliminary efficacy in improving sleep, assessed via a combination of actigraphy, mattress-sensors, sleep diaries (all analyzed daily), as well as pre– and post-BCAA overnight polysomnography for sleep staging, cognition, and quality of life measures. Results indicated high feasibility and acceptability of this fully remote protocol among Veterans with TBI.

## Introduction

Traumatic brain injury (TBI) is common among US Veterans and military service members [1,2] and often complicated by post-concussive sequelae [3,4] that does not resolve in the weeks or months following injury [5]. Although post-concussive sequelae are often highly heterogenous, sleep disturbances are very common [6–10], affecting 50-70% of individuals with TBI [11], and exacerbate other functional outcomes persisting for >20-25 years post-injury [12–15]. Similarly, cognitive disturbances, either independent of or secondary to sleep disturbances, are also highly prevalent (60-80%) in the sub-acute/chronic phase of recovery from TBI [16,17]. Collectively, the economic impact of TBI-related sleep disorders is staggering [18], especially once considering sleep-related exacerbation of post-concussive symptoms, cognition, neuropsychiatric function, and chronic pain, among others [19–22], all of which may delay or completely prevent return to the workforce, community integration, and family life [23].

Therapeutic approaches to improve sleep in patients with TBI are limited to either being symptom directed, pharmacologic, or with substantial patient/provider burden, often compounded by marginal efficacy and poor patient acceptability. Furthermore, treatment for related post-concussive symptoms can complicate the overall clinical picture, for example, opioids, anti-depressants, or anxiolytics may be clinically warranted for symptomatic relief, but indirectly contribute to sleep impairments (i.e., suppressing slow-wave or rapid eye movement sleep, exacerbating sleep-disordered breathing, etc.). Indeed, mechanistic-based, targeted approaches with minimal side effects are desperately needed.

Prior preclinical work over the past 15 years has identified dietary supplementation with branched chain amino acids (BCAA: isoleucine, leucine, and valine) to have ameliorated both sleep and cognitive impairments in a rodent model of TBI [24,25]. Mechanistically, these improvements were likely driven by BCAAs restoration of glutamate within presynaptic nerve terminals contacting orexin neurons in the hypothalamus [26]. Indeed, BCAAs served multiple physiologic roles, however, within the CNS these amino acids provided substrate for the majority of *de novo* glutamate and thereby GABA (i.e., the main excitatory and inhibitory neurotransmitters, respectively) synthesis [27,28]. In support of BCAA supplementation affecting an overall change in cortical excitation to inhibition (E:I), we have also shown that BCAA supplementation restores E:I balance within the hippocampus and improved hippocampal-dependent memory tasks [25,29]. The efficacy for BCAA supplementation to produce changes in cortical E:I balance was critically dependent on consuming these amino acids in isolation, rather than in combination with a high concentration of other amino acids (i.e., via a mixed meal). The majority of BCAAs are not metabolized by the liver and instead freely pass into the systemic circulation, which causes plasma concentrations to rise rapidly after ingestion and in proportion to their dose (or content via solid food) [30]. Plasma BCAAs gain entry to the CNS interstitial fluid and CSF by way of blood brain barrier endothelial cell transporters [31–33], which are non-specific and move amino acids according to their relative concentration gradients. Therefore, competing plasma amino acids (e.g., tryptophan, tyrosine, and phenylalanine) will limit CNS BCAA uptake and provide additional substrate for potentially competing monoamines and catecholamines in the CNS. Finally, our preclinical work has tested the optimal dosing, duration, and route of administration of BCAA effects in mice [29]. Similarly, there has been substantial data in humans demonstrating promising long-term safety profiles for BCAA supplementation, with evidence for no adverse effects following multiple years of supplementation [27,34,35], complemented by well described anti-catabolic and even anti-nociceptive effects [36,37] through investigation in a multitude of physiologic domains (e.g., exercise physiology, hepatic pathology, and various neurological and psychiatric disorders [27,38–54]).

Taken together, there is compelling scientific precedent, mechanistic rationale, and human safety data to support the testing of BCAA supplementation in humans with TBI. Recent work by our group has initiated this translation, demonstrating preliminary feasibility, acceptability, and limited efficacy for BCAA supplementation to improve sleep in 1) Veterans in the chronic phase of recovery from TBI (NCT03990909) [55], and 2) adolescents in the acute phase of recovery from TBI, i.e., the HIT-HEADS randomized clinical trial (NCT0186040) [56]. However, the next step in this translation is demonstrating efficacy through a full-scale double-blind placebo-controlled randomized clinical trial.

The present manuscript describes the protocol we employ in the context of Veterans in the chronic phase of recovery from TBI, which notably, is a fully remote format administered within the Department of Veterans Affairs (VA). This is a challenging population to study, not only due to significant participant disability/burden, but also due to the logistical and regulatory challenges of cloud-based remote studies within the VA. This randomized clinical trial, SmART-TBI (supplementation with amino acid rehabilitative therapy in TBI; NCT04603443), proposed to enroll n = 120 Veterans allocated 1:1:1:1, to one of three BCAA doses (high, 30g b.i.d.; medium, 20g b.i.d.; and low, 10g b.i.d.) and one placebo-control (rice protein, 10g b.i.d.) over a 26-week duration spanning 2-weeks of baseline, 12-weeks of intervention, and 12-weeks of follow-up. Both investigators and participants were blinded to which condition they were randomized to, and thus, all participants engaged with study investigators to the same extent and with the same degree of expectancy. Here we present a detailed description of the clinical trial design, execution, interim feasibility/acceptability measures of the remote protocol, and the statistical analysis plan for future efficacy analyses once data collection is completed.

## Materials and methods

### Overview and ethics

SmART-TBI is a double-blind placebo-controlled randomized clinical trial (NCT04603443), investigating, primarily, the feasibility and acceptability of implementing a fully remote clinical trial within the VA, and secondarily, efficacy for dietary BCAA supplementation to improve sleep quality, cognitive function, and quality of life in Veterans with TBI. Participants were randomized via co-variate adaptive randomization controlling for age, sex, TBI recency, pain, depression, and PTSD in a 1:1:1:1 allocation ratio across one of three BCAA doses (high, 30g b.i.d.; medium, 20g b.i.d.; and low, 10g b.i.d.) and one placebo-control (rice protein, 10g b.i.d.) over a 26-week duration spanning 2-weeks of baseline, 12-weeks of intervention, and 12-weeks of follow-up. All participants expressing interest and who screened eligible, were enrolled in the study according to the Standard Protocol Items and Recommendations for Interventional Trials (SPIRIT) guidelines (**Fig 1**).

**Figure 1.**
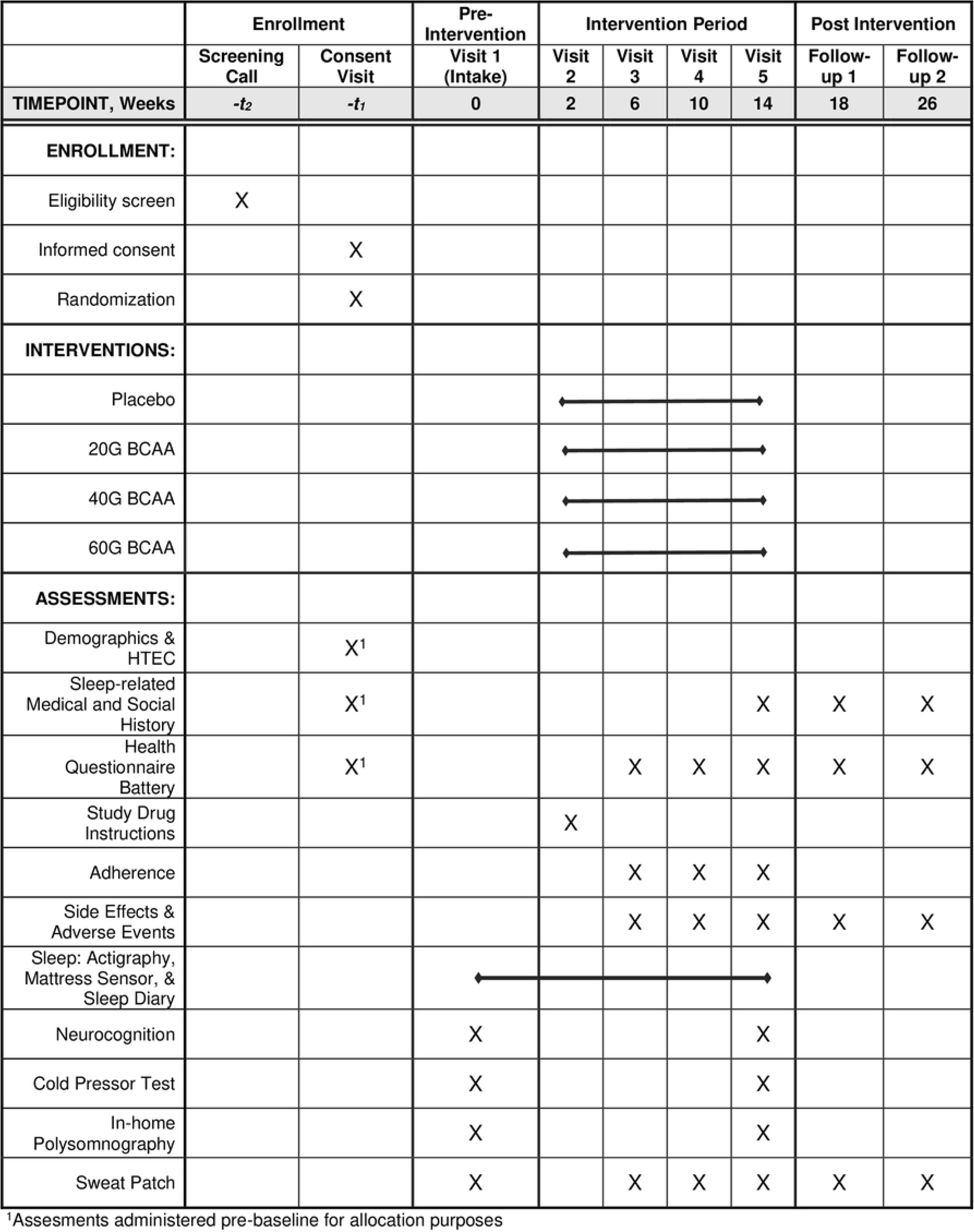
SPIRT outcomes across time. The Standard Protocol Items: Recommendations for Interventional Trials (SPIRIT) timeline for SmART-TBI.

This project was sponsored by the VA and approved by a joint institutional review board (IRB) at the VA Portland Health Care System (VAPORHCS; #4413) and Oregon Health & Science University (OHSU; #21273). All participants provided verbal and written informed consent prior to participation.

### Study Setting

Research staff operated out of the VA Portland Healthcare System (VAPORHCS) in Portland, OR, USA although data collection was primarily carried out in participants homes given the remote nature of this protocol. Participants were able to attend virtual visits from any location within the United States with adequate cellular service, Wi-fi connection, and privacy to communicate with study coordinators and complete study tasks. However, participants were also offered an in-person option at VAPORHCS.

### Sample Size

The sample size was calculated to support the analysis of the primary outcomes, feasibility and acceptability. Completion of the study was defined as completion of at least the 2-week baseline period and 4 weeks of intervention, with a primary outcome being completion of the Insomnia Severity Index (ISI) at both time points.

The target sample size for enrollment is n = 120 (n = 30 per group), which provides 80% power to discern 75% differential retention. 75% retention will leave n = 104 completed participants (n = 26 per group) which provides 85% power to discern 80% differential adherence. Thus far, n = 127 participants have been randomized, n = 15 are currently active in the baseline period, n = 94 have started the intervention, and n = 18 withdrew. Of these n = 94 participants who have started the intervention, n = 87 have met the minimum requirements needed to be considered complete, and n = 7 are currently active during the first 4-week intervention period.

Due to this studies covariate adaptive randomization scheme (see below), there is the potential to exceed this study’s target enrollment/completion goals to ensure balanced and minimum group number requirements. We anticipate enrollment will be completed mid-2025, with data analysis and dissemination of results by end-2025.

### Recruitment

Participants were recruited nationwide, with additional local efforts from flyers and clinician referrals within the VAPORHCS, OHSU, and surrounding Portland, Oregon metropolitan (**Fig 2**). The most successful recruitment methods included repositories, referrals, internet-based advertisements, flyers, and radio ads. All are described in more detail below.

**Figure 2.**
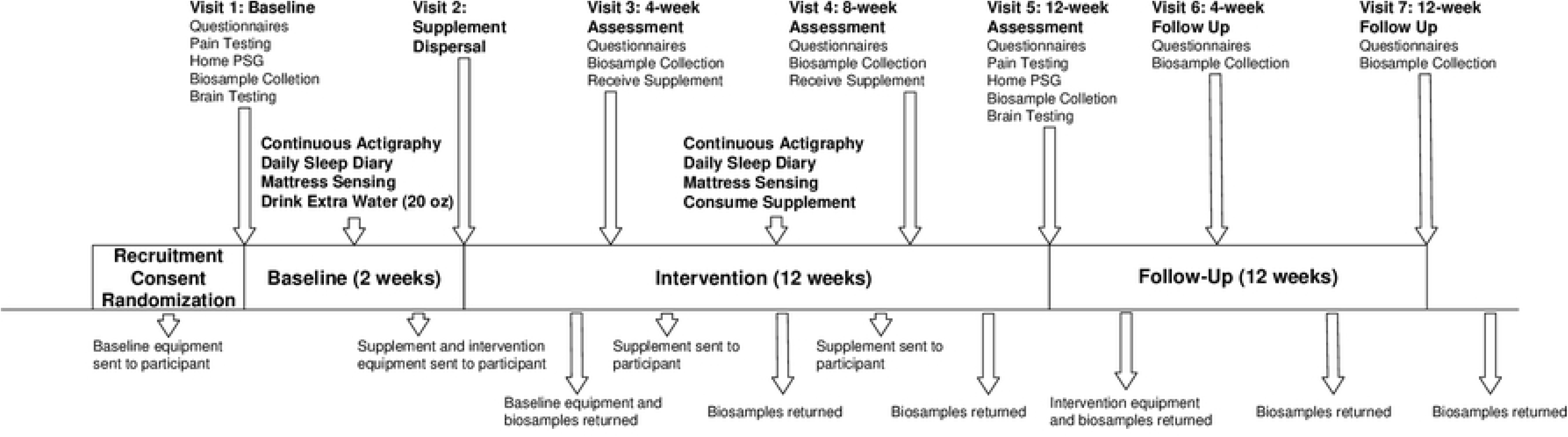
Participant flow chart. Diagram illustrating steps involved with SmART-TBI, including when participant study visits are held, which outcomes are collected when, the duration of each study period as well as when equipment and biospecimens are shipped to/from participants place of residence.

### Repositories

Participant repositories developed from prior and ongoing studies within the VAPORHCS were utilized, which collectively include ∼1500 Veterans who have consented to being recontacted for future research opportunities. Regulatory approvals were in place to share identifiable data and participant contact information with approved SmART-TBI study team personnel. Additionally, we leveraged the VA Informatics and Computing Infrastructure (VINCI), for access to broad VA data and resources while maintaining Veterans’ privacy and confidentiality. Study staff mailed opt out letters to Veterans in the VINCI system describing the SmART-TBI study. If opt out responses were not received within 2 weeks, contact was initiated for eligibility assessment. ResearchMatch.org was also used to refer and connect participants to our project; interested participants created profiles with eligibility criteria and were connected to studies that matched their profiles.

### Referrals

Multiple routes of participant referrals are utilized. First, clinicians within the VAPORHCS and associated community outpatient clinics, as well as OHSU may refer participants for further eligibility assessment. Second, ongoing studies within the Sleep & Health Applied Research Program referred participants who either did not qualify for other studies or have completed said studies. Third, ongoing work sponsored by the Department of Defense Military Traumatic Brain Injury Initiative (MTBI^2^) provided a referral service for those interested in TBI research opportunities, called the TBI Research Opportunities and Outreach for Participation in Studies (TROOPS) program. A TROOPS referral coordinator provided contact information of eligible and interested TROOPS participants via VA-authorized end-to-end encrypted email for all messages containing PHI/PII.

### Website advertisement

Website advertisements were used to direct interested participants to our study. This includes advertisements within the overall VAPORHCS website, as well as affiliated centers and labs (e.g., the VA National Center for Rehabilitative Auditory Research. Similarly, OHSU maintains a research opportunity website where our project is advertised, which is also pushed to the OHSU Brain Institute. Lastly, our research program’s website (www.sharplabpdx.com) describes current research opportunities and instructions on how to join.

### Flyers, social media, and radio

Study fliers were placed in the VAPORHCS primary clinic elevators, the Salem VA Clinic, and the Vancouver VA Medical Center. These flyers were also shared with local clinicians, study sponsors and other partners for broad dissemination. Social media avenues include leveraging existing blogs that directly advertise for SmART-TBI and other sponsored projects, which contributes participants to the overall TROOPS referral program; Craigslist advertisements; GovDelivery, a marketing subscription email service; and other VA-approved accounts. IRB-approved visual banners and audio ads aired on local AM radio stations were also utilized to recruit participants within the Portland-metro area.

### Eligibility criteria

Those eligible to enroll in this study must have incurred at least one TBI, be Veterans over 18 years of age located within the United States, have a stable status on other pharmacological and/or behavioral sleep interventions, and have the ability to provide informed consent and comply with the study protocol. If a potential participant has recently started a new sleep intervention at the time of screening/consent, they must wait for their intervention to stabilize before moving forward in the study. Due to the remote nature of the study, participants must also have a reliable mailing address, email address, phone number, and Wi-fi connection. See **Table 1** for the interim analysis demographics.

**Table 1.** Demographic data for participants that have completed baseline and started the intervention.

|  | Placebo<br>(n=24) | Low<br>(n=20) | Medium<br>(n=21) | High<br>(n=29) |
| --- | --- | --- | --- | --- |
| **Age, years** | 47±10.4 | 59.6±9.8 | 46.2±10 | 50.8±9.8 |
| **Education** |  |  |  |  |
| Highschool/GED | 0 (0.0%) | 1 (5%) | 1 (5%) | 0 (0%) |
| Associates/Some College | 14 (58%) | 10 (50%) | 10 (48%) | 11 (38%) |
| Bachelor's Degree | 5 (21%) | 4 (20%) | 6 (29%) | 6 (21%) |
| Master's Degree | 4 (17%) | 3 (15%) | 3 (14%) | 10 (35%) |
| Doctorate | 1 (4%) | 2 (10%) | 1 (5%) | 2 (7%) |
| **Sex** |  |  |  |  |
| Male | 16 (67%) | 13 (65%) | 15 (71%) | 19 (66%) |
| Female | 8 (33%) | 7 (35%) | 6 (39%) | 10 (34%) |
| **Race and Ethnicity** |  |  |  |  |
| Ethnicity, *Hispanic/Latine* | 2 (8%) | 2 (10%) | 2 (10%) | 3 (10%) |
| Race, *White* | 21 (87%) | 17 (85%) | 17 (81%) | 24 (83%) |
| Race, *Asian* | 1 (4%) | 1 (5%) | 1 (5%) | 0 (0%) |
| Race, *Black of African American* | 3 (12%) | 0 (0%) | 2 (10%) | 4 (14%) |
| Race, *American Indian or Alaskan Native* | 1 (4%) | 1 (5%) | 1 (5%) | 0 (0%) |
| Race, *Two or More Races* | 2 (8%) | 1 (5%) | 0 (0%) | 2 (7%) |
| Race, *Other* | 1 (4%) | 1 (5%) | 0 (0%) | 1 (3%) |
| **Participant Residence Time Zone** |  |  |  |  |
| Pacific Standard Time | 18 (75%) | 16 (80%) | 18 (86%) | 20 (69%) |
| Local Portland-Vancouver metro area | 12 (50%) | 12 (60%) | 15 (72%) | 14 (48%) |
| Mountain Standard Time | 0 (0%) | 1 (5%) | 0 (0%) | 0 (0%) |
| Central Standard Time | 5 (20.8%) | 2 (10%) | 1 (5%) | 4 (14%) |
| Eastern Standard Time | 1 (4.2%) | 1 (5%) | 2 (10%) | 5 (17%) |
| **Recruitment Method** |  |  |  |  |
| Repository | 7 (29%) | 5 (25%) | 9 (43%) | 9 (31%) |
| Referral | 3 (12%) | 2 (10%) | 3 (14%) | 5 (17) |
| Website | 4 (17%) | 2 (10%) | 1 (5%) | 5 (17%) |
| Flyers, social media, and radio | 10 (42%) | 11 (55%) | 8 (38%) | 10 (34%) |

Data are presented as mean ± standard deviation, or *n* (%). Group characteristics were balanced via intake and randomization (inverse probability of treatment weighting). However, continuous variables were analyzed via a one-way ANOVA and categorical variables via chi-square.

History of TBI was confirmed through a structured clinical interview using the Head Trauma Events Characteristics form (HTEC; **Table 2**). The HTEC begins with a standard screening question followed by branching logic questions addressing injury type, location, intracranial injury/skull fracture, duration of loss of consciousness (LOC), and anterograde or retrograde post-traumatic amnesia (PTA).

**Table 2.**
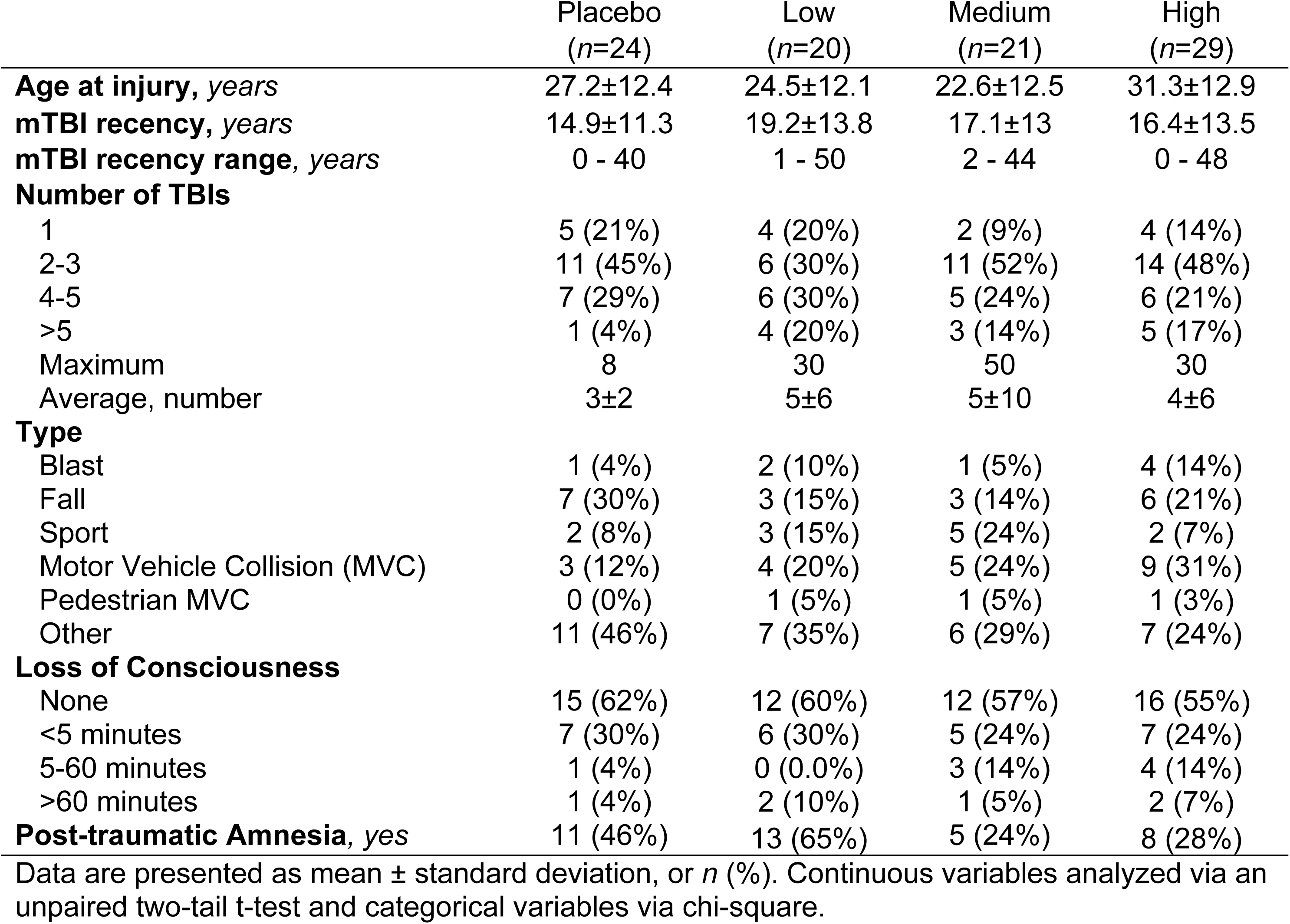
HTEC determined TBI characteristics.

|  | Placebo<br>(*n*=24) | Low<br>(*n*=20) | Medium<br>(*n*=21) | High<br>(*n*=29) |
| --- | --- | --- | --- | --- |
| **Age at injury, years** | 27.2±12.4 | 24.5±12.1 | 22.6±12.5 | 31.3±12.9 |
| **mTBI recency, years** | 14.9±11.3 | 19.2±13.8 | 17.1±13 | 16.4±13.5 |
| **mTBI recency range, years** | 0 - 40 | 1 - 50 | 2 - 44 | 0 - 48 |
| **Number of TBIs** |  |  |  |  |
| 1 | 5 (21%) | 4 (20%) | 2 (9%) | 4 (14%) |
| 2-3 | 11 (45%) | 6 (30%) | 11 (52%) | 14 (48%) |
| 4-5 | 7 (29%) | 6 (30%) | 5 (24%) | 6 (21%) |
| >5 | 1 (4%) | 4 (20%) | 3 (14%) | 5 (17%) |
| Maximum | 8 | 30 | 50 | 30 |
| Average, number | 3±2 | 5±6 | 5±10 | 4±6 |
| **Type** |  |  |  |  |
| Blast | 1 (4%) | 2 (10%) | 1 (5%) | 4 (14%) |
| Fall | 7 (30%) | 3 (15%) | 3 (14%) | 6 (21%) |
| Sport | 2 (8%) | 3 (15%) | 5 (24%) | 2 (7%) |
| Motor Vehicle Collision (MVC) | 3 (12%) | 4 (20%) | 5 (24%) | 9 (31%) |
| Pedestrian MVC | 0 (0%) | 1 (5%) | 1 (5%) | 1 (3%) |
| Other | 11 (46%) | 7 (35%) | 6 (29%) | 7 (24%) |
| **Loss of Consciousness** |  |  |  |  |
| None | 15 (62%) | 12 (60%) | 12 (57%) | 16 (55%) |
| <5 minutes | 7 (30%) | 6 (30%) | 5 (24%) | 7 (24%) |
| 5-60 minutes | 1 (4%) | 0 (0.0%) | 3 (14%) | 4 (14%) |
| >60 minutes | 1 (4%) | 2 (10%) | 1 (5%) | 2 (7%) |
| **Post-traumatic Amnesia, yes** | 11 (46%) | 13 (65%) | 5 (24%) | 8 (28%) |

Data are presented as mean ± standard deviation, or *n* (%). Continuous variables analyzed via an unpaired two-tail t-test and categorical variables via chi-square.

Those eligible to enroll in this study cannot have a diagnosis of Maple Syrup Urine Disease or Amyotrophic Lateral Sclerosis, work third shift (e.g., 2300 – 0700 or equivalent) or any number of days overnight, cannot consume BCAAs unrelated to the study supplement, and cannot be pregnant or trying to become pregnant for the duration of their participation in the study. If a potential participant is consuming a BCAA supplement at the time of screening/consent, they must cease consumption of BCAAs for at least two weeks before continuing in the study.

### Screening and informed consent

Participants were screened for eligibility either in-person or over the phone following an IRB-approved script. Depending on the recruitment route, participants may first complete a “pre-screening” online survey via REDCap, hosted by the OHSU Oregon Clinical & Translational Research Institute. All participants who complete this pre-screening survey were contacted via phone to complete follow-up screening. Once a participant is screened over the phone, they are either consented, informed of their ineligibility, or marked as uninterested.

Informed consent was completed over the phone or via video conference with the participant and a study coordinator according to standard practice, which includes an addendum for permission to store their data and biological samples for future analyses and studies. The study protocol was approved to also mail consent documentation to participants’ residences or to obtain informed consent using the VA-approved signing service DocuSign (www.docusign.com). DocuSign is an e-signature service that hosts the informed consent document and HIPAA authorization for digital signatures, enabling same day screening and consenting, and was introduced to the VA system in March 2021. SmART-TBI was one of the first studies to successfully adopt and implement DocuSign within our local VA, which occurred in July 2021. Digital delivery of documentation is sent using DocuSign’s encryption service with a completed fully signed copy sent to both parties. Local storage of this documentation is housed on the secure fire-wall protected VA Research Drive. Significant advantages exist using DocuSign over physically mailing documentation back and forth, including a substantial time savings and minimization of errors (e.g., missing signature), which only adds further delays. One caveat is that DocuSign does not adjust time of day when signing parties are in different time zones. In these cases, an additional Note to File was included to explain any large discrepancies in timestamps between consenters and participants.

### Randomization, allocation, and masking

After receiving informed consent and HIPAA authorization, participants were randomized in a 1:1:1:1 allocation ratio and recorded according to the Consolidated Standards of Reporting Trials (CONSORT; **Fig 3**).

**Figure 3.**
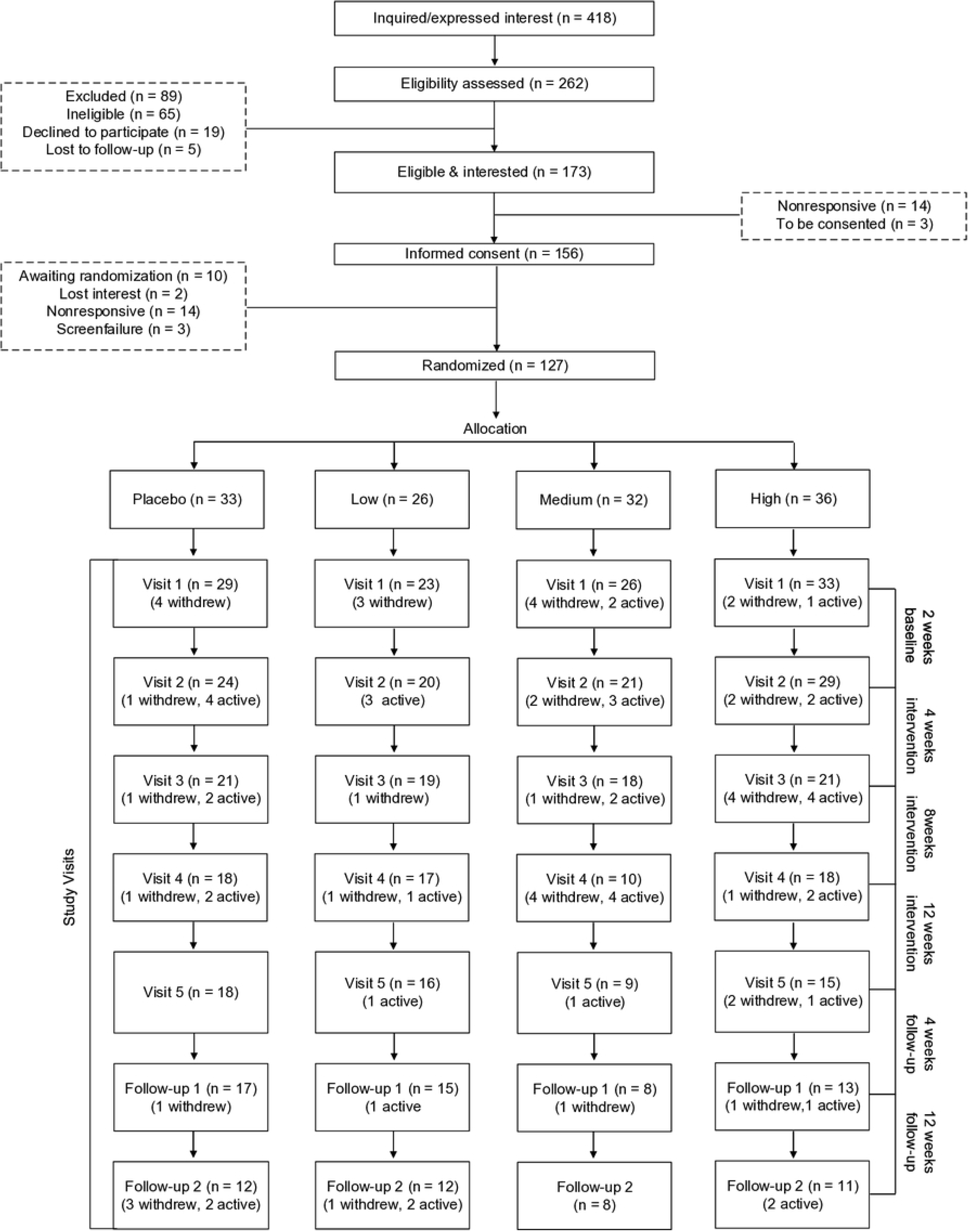
CONSORT diagram. The Consolidated Standards of Reporting Trails (CONSORT) diagram for participants in SmART-TBI to date.

Randomization was achieved via co-variate adaptive methodology which controls *a priori* for age, sex, TBI recency, pain level, depression, and PTSD. To ensure research staff were blinded, information gathered at the baseline visit was provided to the study biostatistician, who then randomized participants using an algorithm unknown to study coordinators. This protocol also maintained the double-mask between investigators and participants by leveraging our local VA Research Pharmacy to coordinate the packing and distributing of dietary supplements, using discreet/uniform packing across groups. Apart from the pharmacy, only the biostatistician and one external lab member (not a member of the study team) were aware of the treatment group doses, for allocation and emergency purposes respectively; neither of which interacted with participants in any capacity. A participant was only unmasked if undergoing a medical emergency in which information about their supplement dose was necessary for them to receive proper treatment. If such a case occurred, the study’s responsible clinician of record was unmasked and study coordinators ceased all contact with the participant, to remain masked to treatment groups.

## Interventions

### Dietary supplement

This protocol used three different dosing parameters for BCAAs in the active arm and one placebo-control condition. All groups followed the same guidelines of consuming one packet of supplement mixed with 20 oz of water within 2 hours of waking, and again ∼6-8 hours later. Participants were provided shaker bottles (BlenderBottle, Trove Brands, LLC, Lehi, UT, USA) and were advised to consume the drink over the course of an hour to avoid digestive discomfort.

With the active BCAA group, there were low (10 g b.i.d.), medium (20 g b.i.d), and high (30 g b.i.d) doses. All BCAA packets used the same commercially available BCAA (BulkSupplements, Henderson, NV, USA) in a 2:1:1 ratio for Leucine, Isoleucine and Valine. Thus, the low, medium, and high dose groups consumed a daily total of 10/5/5 g, 20/10/10 g, or 30/15/15 g of Leucine/Isoleucine/Valine, respectively.

The placebo-control was 10 g b.i.d. of rice protein (NutriBiotic Certified Organic Rice Protein Plain). Rice protein is naturally low in BCAA concentration at ∼6.8% Leucine, ∼3.4% Isoleucine, and ∼6% Valine. Participants in this group are therefore consuming a daily total of ∼1.36 g/0.68 g/1.2 g of Leucine/Isoleucine/Valine, respectively. As described earlier, when consumed with a full spectrum of other amino acids the ability for BCAAs to cross the blood brain barrier is further reduced. Thus, this placebo condition controls for protein intake with minimal BCAA effects.

### Fidelity adherence monitoring

Adherence to the study intervention was monitored in three ways. First, participants self-report their usage daily as part of their sleep diary (described below in *Outcomes*). Second, participants report the number of unused supplement packets they have left at every check-in visit and return all unused packets via mail at the end of the intervention period. Third, sweat-based biospecimen samples (sweat patch; PharmChek Fort Worth, TX, USA) will be assayed for BCAA concentrations on study close-out. Previous pilot work leveraged human plasma samples to demonstrate a significant increase in plasma BCAA concentration in the BCAA group [55], however, consistent with the present clinical trials fully remote format, this assay was adapted for sweat. Samples were collected over a 24-hr period at the beginning of baseline, after 4-, 8– and 12-weeks of intervention, and at 4– and 12-weeks post-treatment. Thus, this assays for a relative increase in total BCAA concentration compared to baseline, which is expected to persist through intervention periods and decline during follow-up.

## Outcomes and data collection

### Primary: Feasibility and Acceptability

The primary outcome in SmART-TBI was demonstrating overall feasibility and acceptability for BCAA supplementation in Veterans with TBI. Protocol feasibility was demonstrated by way of participant retention whereas acceptability was demonstrated through protocol adherence. The threshold for meeting adequate participant retention and adherence was ≥70% (described below).

Adherence was measured (see *Fidelity adherence monitoring*) for the entire 12-week supplement period and analyzed in 4-week segments to examine between-group differences. Adherence was defined as the consumption of at least one supplement packet per day at least five days a week, equaling a minimum of 20 packets consumed per 4-week period. Retention, defined as having completed the preceding treatment and study tasks due at that time point, was retrospectively calculated for Visits 2-7 and analyzed for between-group differences. Data completeness was measured across all time points in the study and retrospectively calculated based on the periodic and continuous data used to monitor the other primary outcomes. The biological samples (retention: completion of study tasks) and questionnaire batteries (retention: completion of study tasks; adherence: frequency of consumption, side effects, protocol deviations) were collected periodically at every study visit (excluding visit 2) and marked as complete or incomplete based on their presence in study databases and freezer storage. Daily sleep diaries (retention: completion of study tasks; adherence: frequency of consumption) were collected continuously from the start of baseline to the end of the supplement period. For the baseline period, data completeness of the sleep diaries was defined as the presence of at least five consecutive days of diary responses at any point during the baseline period. For each of the 4-week supplement period segments, data completeness was defined as the presence of at least five consecutive days of diary responses in the last two weeks of the segment.

### Exploratory: Preliminary efficacy for sleep

Potential changes in sleep will be evaluated via validated self-report measures (e.g., Insomnia Severity Index, ISI [57]; Epworth Sleepiness Scale, ESS [58]; Functional Outcomes of Sleep Questionnaire-10, FOSQ-10 [59]; Sleep Hygiene Index, SHI [60]), and objective metrics derived from wrist-based actigraphy (Philips Respironics, Bend, OR, USA), mattress sensors (Emfit QS, Vaajakoski, Finland), and home-based polysomnography (Somnomedics, Randersacker, Germany). Our primary comparison is baseline relative to 4-weeks of intervention, however, longitudinal changes and analyses across the 12-week intervention period will be explored.

Daily sleep diaries were collected using the TWILIO™ platform, which directly integrates with our HIPAA compliant secure REDCap database. Participants were prompted in the morning via either SMS or email, first at 8:00 AM with reminders sent at 10:00 AM and 12:00 PM if no response has been entered. If participants do not respond within that 24-hr period, earlier prompts expire ensuring all data is collected within 24 hrs. Sleep diary metrics include self-reported time in bed/asleep, time out of bed/awake, number of awakenings during the night, and (during the intervention period) supplement consumption.

Actigraphy was collected via a solid-state piezoelectric monoaxial accelerometer to measure activity and silicon photodiodes to measure photopic light (5-100,000 lux; 400-900 nm), with data sampled at 32 Hz and aggregated into 2-minute epochs (Philips Respironics). Philips Actiware (v 6.3.0) was used to process and analyze these data relying first on the automated algorithmic detection of rest-active periods, with further refinement based on *a priori* defined outcomes collected in the sleep diary. For example, participants are queried whether or not their sleep was unusually disrupted (ranging from being ill to falling asleep or being awoken at usual times due to things outside of their control), and if so, this 24 hr period was excluded from analysis. Participants were directed to wear the watch on their non-dominant wrist and to keep clothing from covering the light sensor as much as possible.

Emfit mattress sensors use ballistocardiography to record heart rate, respiratory rate, and physical activity of participants, and sleep metrics are generated based on changes in activity levels, similar to actigraphy. Participants were instructed to install the sensor horizontally underneath their mattress at chest level, and a live feed of unidentifiable data was transmitted via cellular network or Wi-fi to an API accessible only to the study team and Emfit representatives. Mattress sensor data was cleaned using the same sleep diary reference method as the actigraphy data.

Overnight polysomnography was collected in participants’ homes (SOMNOmedics, SOMNOtouch). Detailed instructions were provided to participants, and a study coordinator was on-call the night of the recording. Reported data include EEG, EOG, EMG, SpO_2_, heart rate, respiratory rate, thorax and abdomen effort, and audio recording. Data was analyzed using the Domino Light SOMNOmedics software.

### Exploratory: Preliminary efficacy for cognition

Neurocognitive function was evaluated through comprehensive neuropsychological testing at baseline and after 12-weeks of intervention). Completed over video, this assessment was 20-30 minutes in length and evaluates memory, attention, executive function, language and processing speed (e.g., Delis-Kaplan Executive Function System Categorical Verbal Fluency Tests, D-KEFS CVFT; Controlled Oral Word Association Test, COWAT-FAS; Wechsler Adult Intelligence Scale, Fourth Edition, WAIS-IV; Hopkins Verbal Learning Test – Revised, HVLT-R, Benson Complex Figure Test, BCFT). The HVLT-R implements “opal-emerald-pearl” at baseline and “chisel-screwdriver-wrench” post-intervention. This evaluation was also recorded for the accurate transcription of participant responses. Remotely implementing neuropsychological testing precludes administering certain measures (e.g., Trails A and B, Color-Word interference, etc.).

### Exploratory: Preliminary efficacy for self-reported health

Additional relevant outcomes include a broad range of validated self-reported outcomes related to neurobehavioral function, mood, and quality of life (e.g., NIH PROMIS Global Health, Pain Intensity/Interference, and Emotional Distress Anxiety [61,62]; Neurobehavioral Symptom Inventory, NSI [63]; Post-traumatic Stress Disorder Checklist for DSM-5, PCL-5 [64]; Patient Health Questionniare-9, PHQ-9 [65]; World Health Organization Disability Schedule 2.0, WHO-DAS 2.0 [66]). On average, this questionnaire battery requires 30-45 minutes to complete. Questionnaires were sent via email to the participant, and responses automatically imported into REDCap. If questionnaires must be printed and mailed to a participant, a study coordinator manually enters their responses into REDCap with a secondary rater confirming entry accuracy.

### Exploratory: Preliminary efficacy for biomarkers

Sweat patches (PharmChek, Fort Worth, TX, USA) were used to collect biofluid samples, which will be assayed for biomarkers of neuroinflammation and degeneration (e.g., Neurofilament Light Chain, NfL; Glial Fibrillary Acidic Protein, GFAP; Ubiquitin Carboxyl-Terminal Hydrolase L1, UCH-L1; and total Tau, t-Tau), as well as markers of systemic inflammation (e.g., Interleukin-6, IL-6; Interleukin-10, IL-10; and Tumor Necrosis Factor-alpha, TNF-α) [67]. Participants were instructed to affix the sweat patch on their lower abdomen, after cleaning the area with an alcohol swab, and to keep the sweat patch on for 24 hours before storing in their freezer. This sweat patch and two ThermoSafe^®^ U-tek^®^ ice packs are then shipped back to the lab via an insulated PACKIT^®^ freezer bag for storage at –20°C.

### Shipping

This study uses the United States Postal Service (USPS) to ship all packages to and from participants. Small, medium, and large USPS flat rate boxes are used throughout the study, and participants are provided with the shipping materials necessary, including pre-paid labels, to return study equipment and biological samples. Packages are sent with enough time to be delivered by USPS before each study visit (**Fig 2**).

## Data safety, monitoring and auditing

### Harms and AE/SAE reporting

It is possible that participants may experience mild GI discomfort after taking the supplement, discomfort from the sweat patch adhesive, polysomnography electrode adhesive, or actiwatch, and possible psychological discomfort from questionnaire items regarding suicidality, depression, traumatic brain injury, PTSD, and prior military experience. Adverse events are reported to the VA IRB and the VA research and development department within 24 hours on their discovery, and all adverse events are evaluated by the IRB and the VA Central Data Monitoring Committee (DMC) yearly.

### Confidentiality

All PHI/PII used in or obtained from recruitment was done so with consent from the pertaining individual and was exclusively managed by HIPAA-authorized study coordinators. All physical copies of study materials were stored in a locked file cabinet, within a locked room, behind secure-entry badge access doors within VAPORHCS. Study personnel refer to participants and their data according to their coded identification number, in verbal and written communication, unless done so on a secure VA channel. All main lists or main keys to the coding system remain secured on the VAPORHCS Research drive, behind the VA firewall and accessible only to approved study team members. Participant data is also stored on the HIPAA-secure REDCap database, hosted on OHSU secure servers, maintained by OHSU Oregon Clinical and Translational Research Institute, and accessible only to approved study team members. Data sharing may occur following approval of joint institutional Data Use Agreements which specify whether data is deidentified, coded, or identifiable.

The OHSU Oregon Clinical and Translational Research Institute hosts and manages data security and backup of the HIPAA compliant REDCap database, where participant response data is stored. They have checks and balances coded into the database to allow for proper data protection, historical information and qualitative notes to be maintained. Access is protected via OHSU login information and user credentials once the database is shared from previously approved study personnel. This database is HIPAA-secure and all those interacting with the database are trained and knowledgeable of HIPAA and VA privacy policy.

### Monitoring

The centralized Data Monitoring Committee (DMC) of the VA Clinical Science Research & Development (CSR&D) provides independent oversight on the safety, conduct, and integrity of the project through evaluation of participant accrual and retention, adverse events, and data analyses. Reports on the safety and status of the study were submitted to the DMC annually, and meetings are held by the DMC bi-annually which inform CSRD to recommend conditional or unconditional continuation, probation, or termination.

### Statistical analyses

Statistical analyses will be performed using GraphPad Prism v9 or R, with alpha defined *a priori* at 0.05. Prior to evaluating outcomes, we will perform a thorough descriptive analysis of participant baseline characteristics overall and across study arms. Categorical variables will be described using frequencies and percentages. Histograms and boxplots will be used to assess the distribution of continuous variables. Continuous variables that follow an approximately normal distribution will be summarized using means and standard deviations; skewed continuous variables will be reported as medians and interquartile ranges. We will examine patterns of missingness in outcomes over time, at each visit, and by study arm. As part of our descriptive analyses, we will compare participants who complete versus dropout of the study by baseline characteristics and study arm to ascertain potential biases that may impact this and future studies. Finally, we will assess the characteristics of individuals that do and do not participate. Comparisons between groups will be made using chi-square tests or Fisher’s Exact Tests for categorical variables and T-tests or Mann-Whitney U tests for continuous variables, depending on the characteristics of the data.

Feasibility and acceptability outcomes will be assessed descriptively. Recruitment, participation, retention, and screening failure rates will be reported in a CONSORT flow diagram along with reasons for participation, non-participation, and withdrawal. Adherence to treatment will be measured as the proportion of study drug consumed based on the number of used packets returned to the VA Research Pharmacy and summarized overall and by study arm. Patient satisfaction and reasons with non-adherence will be measured with questions on a five-point Likert scale. Results will be presented graphically as diverging stacked bar charts and summarized using medians and IQR. The number and severity of side effects will be recorded overall and by treatment arm using the Monitoring of Side Effects Scale (MOSES). Total MOSES score as well as GI, neurological, and psychiatric side effect subscales will be collected at each visit during the intervention period.

Inferential analyses will be performed with limited efficacy outcomes in mind. We will use a rigorous set of analyses to explore the magnitude of treatment effects and investigate the effect of BCAA supplementation over time. These results will inform our design of a future trial including selection of a primary outcome. Analyses will follow an intention-to-treat design whereby all patients will be analyzed according to the treatment group to which they were assigned, whether or not they completed the intervention for that group. For these analyses we will begin by assessing the normality of outcome variables and transforming skewed distributions to meet normality assumptions where appropriate. We will also construct plots to graphically inspect each outcome over time, including both graphs of the mean response as well as individual participants’ outcomes for dichotomous treatment groups (placebo and active) and by study arm. To the extent that we are able, we will graphically assess outcomes in subgroups defined by TBI recency, pain level, OSA, depression, PTSD, and insomnia and compare them to the full sample.

We will investigate the effect of BCAA supplementation for the sleep outcomes outlined in **Table 3** using general linear mixed-effects regression. The dependent variable in the model will be individual sleep outcomes. Treatment group (placebo and active groups), weeks from baseline, baseline outcome score, baseline characteristics used in randomization (age, sex, TBI recency, pain level, depression, and PTSD), and treatment-by-week interaction will be included in the regression as fixed effects. A random intercept term for participants will address the correlation among measurements from the same individual. We will maximize power by (1) considering weeks from baseline as a continuous variable and (2) including the randomization variables in the model. From this model, we will estimate the treatment effect with corresponding 95% confidence intervals (CI) for each visit and present them graphically. The treatment-by-week interaction will be used to assess whether sleep outcomes differ between treatment groups over the course of the study. Due to the scope of this feasibility study, none of the analyses in Aim 2 will be adjusted for multiple comparisons.

**Table 3.** Retention, adherence, adverse events and Monitoring of Side Effect Scale data.

|  | Placebo<br>(n=24) | Low<br>(n=20) | Medium<br>(n=21) | High<br>(n=29) |
| --- | --- | --- | --- | --- |
| **Participant Retention** |  |  |  |  |
| Baseline | 24/24 (100%) | 20/20 (100%) | 21/21 (100%) | 29/29 (100%) |
| Intervention | 21/22 (95%) | 19/20 (95%) | 18/20 (90%) | 21/25 (84%) |
| **Supplement Adherence** |  |  |  |  |
| Intervention | 20/21 (95%) | 18/19 (95%) | 17/18 (94%) | 19/21 (90%) |
| **Adverse Events** | 11 | 9 | 6 | 10 |
| **Serious Adverse Events** | 1 | 2 | 1 | 4 |
| **MOSES Category** |  |  |  |  |
| Ears/eyes/head/face | 10 (28%) | 4 (12%) | 5 (20%) | 1 (3%) |
| Mouth | 3 (8%) | 3 (9%) | 4 (16%) | 2 (6%) |
| Nose/throat/chest | 10 (28%) | 7 (21%) | 6 (24%) | 9 (29%) |
| Gastrointestinal/digestive | 17 (47%) | 7 (21%) | 6 (24%) | 9 (29%) |
| Musculoskeletal/Neurological | 11 (31%) | 9 (26%) | 5 (20%) | 3 (10%) |
| Skin/body temperature | 10 (28%) | 7 (21%) | 5 (20%) | 7 (23%) |
| Urinary/genital | 4 (11%) | 6 (18%) | 3 (12%) | 2 (6%) |
| Psychological | 15 (42%) | 13 (38%) | 9 (36%) | 8 (26%) |

Data are n (%), analyzed via chi square. MOSES, Monitoring of Side Effects Scale

To address the secondary hypothesis of a dose dependent improvement, we will follow the modeling strategy described above for sleep outcomes using a continuous variable for BCAA dose in lieu of treatment group. Sleep outcomes will be the dependent variable. As before, models will include fixed terms for baseline outcome score, weeks from baseline, and baseline characteristics used in randomization and random intercept term for participants. No interaction term will be included. Using these models, we perform linear tests for trends to assess dose-response at 4, 8, and 12 weeks. We will also estimate and graph the marginal mean response and 95% CI for each dose and visit to provide preliminary data about the effect of BCAA supplementation over time for varying doses.

We will use linear mixed-effects regression as described previously to explore the impact of BCAA supplementation on cognitive and quality of life outcomes. Briefly, individual outcomes will be modeled by treatment group (placebo and active groups), weeks from baseline, baseline outcome score, baseline characteristics used in randomization, and treatment-by-week interaction as fixed effects and participants as a random intercept term. Treatment effects with corresponding 95% CI will be estimated for each visit then presented graphically. The treatment-by-week interaction will be used to assess whether cognitive and quality of life outcomes differ between treatment groups over the course of the study.

Although every effort will be made to minimize the risk of systematic missing data, we recognize that it may occur. Because the presence of missing data can distort study results, we will perform sensitivity analyses to assess the influence of missingness and attribute this to either “Random” or “System” causes. We will use these analyses to evaluate the robustness of our findings and potentially obtain better estimates of the magnitude of effects to inform our next trial. For these analyses, we will assume that missingness is either due to missing at random, or not missing at random (e.g., “System” causes), with the underlying realization that data are rarely missing completely at random. We will follow the recommendations by Jakobsen et al.^97^ to handle missing data for our assessment:

(1) If the proportion of missing data is less than 5% and there is not evidence that certain patient groups have differential patterns of missingness (based on assessments in Aim 1), we will consider the potential impact of missing data to be negligible and impute of “best” and “worst” case scenarios for missing data to estimate plausible ranges of BCAA and placebo effects.
(2) If missingness is less than 25% in the dependent variable, we will implement Markov Chain Monte Carlo (MCMC) multiple imputation to impute missing values then analyze and combine five imputed datasets. MCMC algorithms exist to handle longitudinal patterns of missingness.
(3) In the unlikely event that missingness is extensive, we will not attempt to impute missing values. In this case, the results of the complete case analyses will be reported along with reasons for missingness. In all cases, we will discuss and report the extent of missingness with a clear discussion of study limitations.

## Results

Study enrollment is and progression through this project is illustrated in **Fig 2** with enrollment overtime in **Fig 4**. Proposed project milestones over the 4 year study were described as, “*We will allow 9 months for start-up, equipment purchases, new hires, IRB approval, and personnel training. We will allow 9 months at the end of the study to analyze results and transfer data to the NIH Federal Interagency Traumatic Brain Injury Research (FITBIR) database. This will leave 33 months available to enroll participants. Assuming the project results in the anticipated outcomes, we plan to seek additional funding through the VA Merit Award renewal mechanism to examine additional outcomes, expand to additional sites, and/or explore eventual implementation of health services*.”

**Figure 4.**
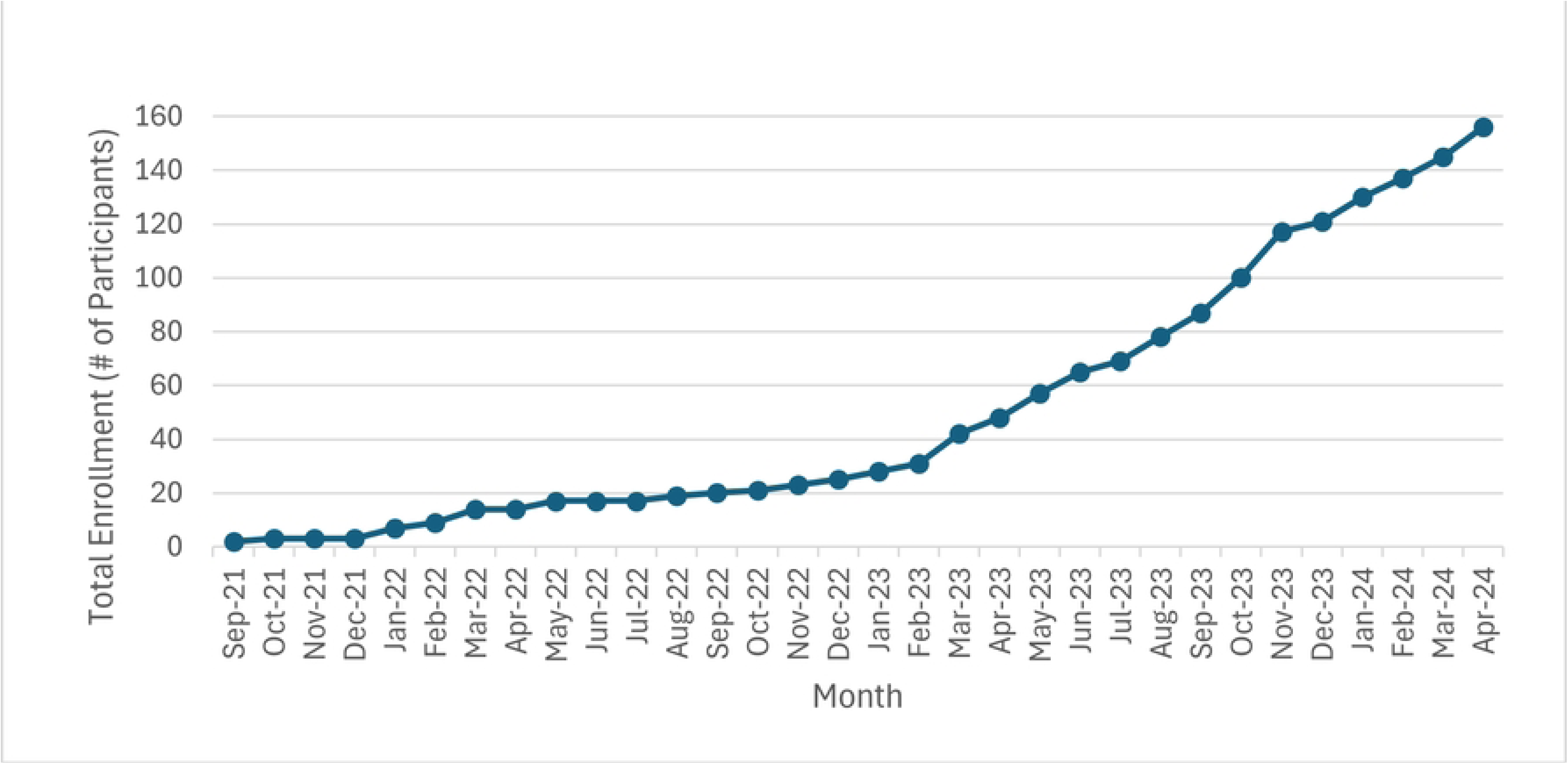
Participant enrollment over time. Visualization of the rate of participant enrollment across this projects performance period to date.

In Year 1 (4/1/21 – 3/31/22), we made excellent progress towards achieving the proposed milestones, including obtaining IRB approval and the necessary protocol modifications required to transition the study to remote procedures in response to restrictions imposed by the COVID-19 pandemic. The prior published pilot study which established the original workflow of SmART-TBI 1.0 was on-site with no remote capabilities [55]. The revised protocol was successfully registered in ClinicalTrials.gov and the VA ART database. Other notable accomplishments in Year 1 included creation and implementation of an entirely new VA workflow for electronic consent (i.e., DocuSign), virtual/electronic administration of screening, questionnaires, and daily sleep diaries, setup of video visits for neuropsychological testing, mailout protocol of all sleep devices including the actiwatch and overnight polysomnography testing, and workflow for the mailout of the BCAA or placebo supplement through the VA mailroom via our VA Research Pharmacy (which was previously distributed on-site). Study-specific flags were also created and implemented through VA VISTA in each patient’s CPRS medical record which alerted treating clinicians about their patient’s participation in this clinical trial. Our first participant was successfully randomized with our new, fully remote protocol in Q3 of 2022. In Year 2 (4/1/22 – 3/31/23), we continued to make strong progress, albeit with slower than anticipated participant enrollment due to continued consequences of the COVID-19 FY22 surge, which impacted both our study staff and the VAPORHCS Research Pharmacy. Due to these factors, only n=25 Veterans were enrolled and randomized in SmART-TBI. We implemented a number of changes to enhance recruitment and workarounds for short staffing.

Subsequently, in Year 3 (4/1/23 – 3/31/24), dramatic improvement was made in getting enrollment and project milestones back on track (**Fig 4**).

Note that inferential analyses for the exploratory preliminary efficacy outcomes from this study will be performed after study close out; however, here we present primary analyses evaluating the feasibility and acceptability of this protocol. Due to the remote format, intervention protocol, and length of the study, it is of value to assess potential attrition, difficulties adhering to the protocol and completing study tasks, and possible barriers to monitoring participant safety during intervention.

Regarding attrition, of the n = 33 participants lost between screening and allocation, n = 28 were terminated due an inability to connect, and of the n = 41 participants lost between baseline and follow-up 2, n = 21 were terminated due an inability to connect (**Fig 3**). However, even with difficulties in communication impeding retention, all four treatment groups exceeded the 70% threshold for retention during the intervention period (**Table 3**). Adherence to the dietary intervention also exceeded the 70% threshold, with all groups at 90% or above and without significant differences in the rates of adverse/serious adverse events. Furthermore, there were no differences detected across groups or time within either the MOSES total score (reflecting an aggregate symptom severity) or individual sub-categories. Of note, the only sub-category *a priori* determined relevant to this intervention was the gastrointestinal/digestive category, and again, no significant differences were observed.

When considering the relative proportions of participants across treatment arms with data for both primary sleep outcomes and exploratory outcomes (**Table 4**), essentially every metric met or exceeds this studies *a priori* defined threshold of 70%. Interestingly, self-reported outcomes during the intervention period for the medium and high dose groups were either modestly sub-threshold (67%) or borderline threshold (71%). Reasons why these were seemingly lower than other, more complicated and burdensome outcomes are unknown though could be attributed to potential technological difficulties

**Table 4.** Proportion of retained participants with data across sleep and exploratory outcomes.

|  | Placebo<br>(n=24) | Low<br>(n=20) | Medium<br>(n=21) | High<br>(n=29) |
| --- | --- | --- | --- | --- |
| **Sleep Outcomes** |  |  |  |  |
| <u>Self-Report</u> |  |  |  |  |
| Baseline | 24/24 (100%) | 20/20 (100%) | 21/21 (100%) | 29/29 (100%) |
| Intervention | 16/21 (76%) | 17/19 (90%) | 12/18 (67%) | 15/21 (71%) |
| <u>Sleep Diary</u> |  |  |  |  |
| Baseline | 22/24 (92%) | 19/20 (95%) | 19/21 (91) | 27/29 (92%) |
| Intervention | 21/21 (100%) | 19/19 (100%) | 17/18 (94%) | 20/21 (95%) |
| <u>Actigraphy</u> |  |  |  |  |
| Baseline | 22/24 (92%) | 19/20 (95%) | 18/21 (86%) | 25/29 (86%) |
| Intervention | 16/21 (76%) | 15/19 (79%) | 13/18 (72%) | 16/21 (76%) |
| <u>Mattress Sensor</u> |  |  |  |  |
| Baseline | 16/24 (67%) | 14/20 (70%) | 14/21 (67%) | 21/29 (72%) |
| Intervention | 20/21 (95%) | 14/19 (74%) | 13/18 (72%) | 18/21 (86%) |
| <u>Polysomnography</u> |  |  |  |  |
| Baseline | 20/24 (83%) | 20/20 (100%) | 21/21 (100%) | 26/29 (90%) |
| Intervention | 14/17 (82%) | 14/16 (88%) | 8/9 (89%) | 12/14 (86%) |
| **Exploratory Outcomes** |  |  |  |  |
| <u>Full Questionnaire Battery</u> |  |  |  |  |
| Baseline | 24/24 (100%) | 20/20 (100%) | 21/21 (100%) | 29/29 (100%) |
| Intervention | 16/21 (76%) | 17/19 (90%) | 12/18 (67%) | 15/21 (71%) |
| <u>Sweat Patch</u> |  |  |  |  |
| Baseline | 23/24 (96%) | 19/20 (95%) | 21/21 (100%) | 25/29 (86%) |
| Intervention | 19/21 (90%) | 15/19 (79%) | 14/18 (78%) | 17/21 (81%) |
| <u>Neurocognitive Assessment</u> |  |  |  |  |
| Baseline | 24/24 (100%) | 20/20 (100%) | 21/21 (100%) | 29/29 (100%) |
| Intervention | 17/17 (100%) | 16/16 (100%) | 9/9 (100%) | 13/14 (93%) |
| <u>Quantitative Pain Testing</u> |  |  |  |  |
| Baseline | 24/24 (100%) | 20/20 (100%) | 21/21 (100%) | 29/29 (100%) |
| Intervention | 15/17 (88%) | 15/16 (94%) | 8/9 (89%) | 12/14 (86%) |
| <u>Oragene</u> |  |  |  |  |
| Baseline | 23/24 (96%) | 18/20 (90%) | 20/21 (95%) | 24/29 (83%) |

Data are n (%), analyzed via chi square.

## Discussion

This represents a detailed description of the SmART-TBI protocol, a placebo-controlled, double-masked randomized clinical trial primarily evaluating the feasibility and acceptability of dietary BCAA supplementation as a potential sleep intervention for Veterans with TBI. Participants were randomized 1:1:1:1 to four groups comprising 3 BCAA doses (30g b.i.d. “High”; 20g b.i.d. “Medium”; and 10g b.i.d. “Low”) and one placebo-control (rice protein, 10g b.i.d.). Supplement was packaged into individual doses by the Portland VA Research Pharmacy, which maintains this study’s double-mask. Outcome measures were assessed following a 2-week baseline period; subsequently after 4 weeks, 8 weeks, and 12 weeks of consumption (i.e., intervention); as well as 4 weeks and 12 weeks after ending consumption (i.e., follow-up). Primary outcomes were feasibility and acceptability of this protocol. Exploratory outcomes included preliminary efficacy measures of sleep quality, cognition, chronic pain, self-reported mental and physical health, quality of life, and sweat-sampled biomarkers implicated in neuroinflammation, systemic inflammation, and neurodegeneration. We have provided interim results on demographics, retention and adherence to the intervention, and the return on data collection methods to show the advantages and disadvantages of remote clinical testing in the VA healthcare system and to demonstrate both the feasibility and acceptability of the current protocol.

Feasibility and acceptability outcomes from the study indicated overall, high feasibility of this fully remote protocol despite the inherent challenges to implementation of randomized clinical trials within a cognitively challenged, highly disabled population such as Veterans with TBI and sleep disruption. Retention and adherence to the supplement ranged from 85-95% across all 4 treatment arms, and no significant differences in adverse events or side effects reported across arms. There were also no significant differences across groups within MOSES categories, and notably, this included the gastrointestinal subscale score. Gastrointestinal discomfort, albeit mild, was not unexpected based on prior pilot data [55,56]. However, the present analysis clearly demonstrates gastrointestinal discomfort from relatively high dose BCAA supplementation does not differ from lower doses or a comparatively benign rice protein control.

This study had a number of strengths, including high rigor in the double-masking, monitoring by an external committee, and implementation of covariate adaptive randomization to weight each treatment group equally for potential confounders such as age, sex, TBI recency, pain, depression, and PTSD. Limitations included a slow study startup complicated by COVID-19 pandemic-imposed restrictions, conversion of the in-person study to a fully remote protocol, prior to regaining momentum after these delays. Other potential limitations include implementing changes to further maximize retention, adherence, and data completeness. Full data analysis and unblinding will occur after study closeout to determine the preliminary efficacy of objective sleep measures, which will be important evidence for biological target engagement.

## Acknowledgements

The authors would like to express their sincere appreciation and gratitude for the participation of our research subjects. Additionally, we would like to thank former/current Sleep & Health Applied Research Program members that have contributed to this line of inquiry (Nadir Balba, PhD; Alisha McBride, RN, BS; Cadence Michel, BS, DC; Allison Keil, BS; Shannon Hardman, BS; Ryan Opel, BS; Peyton Wickham, BS).

## Author contribution

1) Conception and design of the study. JE, AH, AC, ML
2) Acquisition and analysis of data. JE, SS, CO, HC, AE, KP, JB, JD, SH, AH
3) Drafting a significant portion of the manuscript or figures (i.e., a substantial contribution beyond copy editing and approval of the final draft, which is expected of all authors). JE, SS, ML

## Data availability statement

The data underlying this article will be shared on reasonable request to the corresponding author

## References

1. Dewan MC, Rattani A, Gupta S, Baticulon RE, Hung Y-C, Punchak M, et al. Estimating the global incidence of traumatic brain injury. J Neurosurg. 2019;130: 1080–1097. doi:10.3171/2017.10.jns17352

2. Control C for D and. Injury Prevention and Control Traumatic Brain Injury and Concussion. 2016. Available: http://www.cdc.gov/traumaticbraininjury/get_the_facts.html

3. Hoge CW, McGurk D, Thomas JL, Cox AL, Engel CC, Castro CA. Mild Traumatic Brain Injury in U.S. Soldiers Returning from Iraq. New Engl J Medicine. 2008;358: 453–463. doi:10.1056/nejmoa072972

4. Okie S. Traumatic brain injury in the war zone. New England Journal of Medicine. 2005;352: 2043–2047.

5. Rutherford WilliamH, Merrett JohnD, Mcdonali JohnR. Sequelae of Concussion Caused By Minor Head Injuries. Lancet. 1977;309: 1–4. doi:10.1016/s0140-6736(77)91649-x

6. Ouellet M-C, Savard J, Morin CM. Insomnia following Traumatic Brain Injury: A Review. Neurorehab Neural Re. 2004;18: 187–198. doi:10.1177/1545968304271405

7. Ohayon MM, Shapiro CM. Sleep disturbances and psychiatric disorders associated with posttraumatic stress disorder in the general population. Compr Psychiat. 2000;41: 469–478. doi:10.1053/comp.2000.16568

8. Neylan TC, Marmar CR, Metzler TJ, Weiss DS, Zatzick DF, Delucchi KL, et al. Sleep Disturbances in the Vietnam Generation: Findings From a Nationally Representative Sample of Male Vietnam Veterans. Am J Psychiat. 1998;155: 929–933. doi:10.1176/ajp.155.7.929

9. Castriotta RJ, Wilde MC, Lai JM, Atanasov S, Masel BE, Kuna ST. Prevalence and consequences of sleep disorders in traumatic brain injury. J Clin Sleep Medicine Jcsm Official Publ Am Acad Sleep Medicine. 2007;3: 349–56.

10. Duclos C, Dumont M, Wiseman-Hakes C, Arbour C, Mongrain V, Gaudreault P-O, et al. Sleep and wake disturbances following traumatic brain injury. Pathol Biol. 2014;62: 252–261. doi:10.1016/j.patbio.2014.05.014

11. Mathias JL, Alvaro PK. Prevalence of sleep disturbances, disorders, and problems following traumatic brain injury: A meta-analysis. Sleep Med. 2012;13: 898–905. doi:10.1016/j.sleep.2012.04.006

12. Kempf J, Werth E, Kaiser PR, Bassetti CL, Baumann CR. Sleep–wake disturbances 3 years after traumatic brain injury. J Neurology Neurosurg Psychiatry. 2010;81: 1402. doi:10.1136/jnnp.2009.201913

13. Elliott JE, Balba NM, McBride AA, Callahan ML, Street KT, Butler MP, et al. Different Methods for Traumatic Brain Injury Diagnosis Influence Presence and Symptoms of Post-Concussive Syndrome in United States Veterans. J Neurotraum. 2021;38: 3126–3136. doi:10.1089/neu.2021.0031

14. Balba NM, Elliott JE, Weymann KB, Opel RA, Duke JW, Oken BS, et al. Increased Sleep Disturbances and Pain in Veterans With Comorbid Traumatic Brain Injury and Posttraumatic Stress Disorder. J Clin Sleep Med. 2018;14: 1865–1878. doi:10.5664/jcsm.7482

15. Elliott JE, Opel RA, Weymann KB, Chau AQ, Papesh MA, Callahan ML, et al. Sleep Disturbances in Traumatic Brain Injury: Associations With Sensory Sensitivity. J Clin Sleep Med. 2018;14: 1177–1186. doi:10.5664/jcsm.7220

16. Scholten JD, Sayer NA, Vanderploeg RD, Bidelspach DE, Cifu DX. Analysis of US Veterans Health Administration comprehensive evaluations for traumatic brain injury in Operation Enduring Freedom and Operation Iraqi Freedom Veterans. Brain Injury. 2012;26: 1177–1184. doi:10.3109/02699052.2012.661914

17. Vasterling JJ, Proctor SP, Amoroso P, Kane R, Heeren T, White RF. Neuropsychological Outcomes of Army Personnel Following Deployment to the Iraq War. Jama. 2006;296: 519–529. doi:10.1001/jama.296.5.519

18. Colton H, Altevogt B. Sleep disorders and sleep deprivation: an unmet public health problem. HR C, BM A, editors. 2006. doi:10.5860/choice.44-5682

19. Nguyen S, McKay A, Wong D, Rajaratnam SM, Spitz G, Williams G, et al. Cognitive Behavior Therapy to Treat Sleep Disturbance and Fatigue After Traumatic Brain Injury: A Pilot Randomized Controlled Trial. Arch Phys Med Rehab. 2017;98: 1508–1517.e2. doi:10.1016/j.apmr.2017.02.031

20. Lew HL, Otis JD, Tun C, Kerns RD, Clark ME, Cifu DX. Prevalence of chronic pain, posttraumatic stress disorder, and persistent postconcussive symptoms in OIF/OEF veterans: Polytrauma clinical triad. J Rehabilitation Res Dev. 2009;46: 697. doi:10.1682/jrrd.2009.01.0006

21. Rao V, McCann U, Han D, Bergey A, Smith MT. Does acute TBI-related sleep disturbance predict subsequent neuropsychiatric disturbances? Brain Injury. 2013;28: 20–26. doi:10.3109/02699052.2013.847210

22. Worthington AD, Melia Y. Rehabilitation is compromised by arousal and sleep disorders: Results of a survey of rehabilitation centres. Brain Injury. 2006;20: 327–332. doi:10.1080/02699050500488249

23. Corrigan JD, Cuthbert JP, Harrison-Felix C, Whiteneck GG, Bell JM, Miller AC, et al. US Population Estimates of Health and Social Outcomes 5 Years After Rehabilitation for Traumatic Brain Injury. J Head Trauma Rehab. 2014;29: E1–E9. doi:10.1097/htr.0000000000000020

24. Lim MM, Elkind J, Xiong G, Galante R, Zhu J, Zhang L, et al. Dietary Therapy Mitigates Persistent Wake Deficits Caused by Mild Traumatic Brain Injury. Sci Transl Med. 2013;5: 215ra173. doi:10.1126/scitranslmed.3007092

25. Cole JT, Mitala CM, Kundu S, Verma A, Elkind JA, Nissim I, et al. Dietary branched chain amino acids ameliorate injury-induced cognitive impairment. Proc National Acad Sci. 2010;107: 366–371. doi:10.1073/pnas.0910280107

26. Elliott JE, Luche SED, Churchill MJ, Moore C, Cohen AS, Meshul CK, et al. Dietary therapy restores glutamatergic input to orexin/hypocretin neurons after traumatic brain injury in mice. Sleep. 2018;41. doi:10.1093/sleep/zsx212

27. Fernstrom JD. Branched-Chain Amino Acids and Brain function. J Nutrition. 2005;135: 1539S–1546S. doi:10.1093/jn/135.6.1539s

28. Yudkoff M. Brain metabolism of branched-chain amino acids. Glia. 1997;21: 92–98. doi:10.1002/(sici)1098-1136(199709)21:1<92::aid-glia10>3.0.co;2-w

29. Elkind JA, Lim MM, Johnson BN, Palmer CP, Putnam BJ, Kirschen MP, et al. Efficacy, Dosage, and Duration of Action of Branched Chain Amino Acid Therapy for Traumatic Brain Injury. Front Neurol. 2015;6: 73. doi:10.3389/fneur.2015.00073

30. Matthews DE. Observations of Branched-Chain Amino Acid Administration in Humans 1, 2. J Nutr. 2005;135: 1580S–1584S. doi:10.1093/jn/135.6.1580s

31. Smith QR, Momma S, Aoyagi M, Rapoport SI. Kinetics of Neutral Amino Acid Transport Across the Blood-Brain Barrier. J Neurochem. 1987;49: 1651–1658. doi:10.1111/j.1471-4159.1987.tb01039.x

32. Robertson CS, Clifton GL, Grossman RG, Ou C-N, Goodman JC, Borum P, et al. Alterations in Cerebral Availability of Metabolic Substrates after Severe Head Injury. J Trauma Inj Infect Critical Care. 1988;28: 1523–1532. doi:10.1097/00005373-198811000-00002

33. Hawkins RA, O’Kane RL, Simpson IA, Viña JR. Structure of the Blood–Brain Barrier and Its Role in the Transport of Amino Acids. J Nutr. 2006;136: 218S–226S. doi:10.1093/jn/136.1.218s

34. Fernstrom JD. Branched-Chain Amino Acids and Brain Function. American Society for Nutritional Sciences. 2005; 1547–1552.

35. Knapik JJ, Steelman RA, Hoedebecke SS, Austin KG, Farina EK, Lieberman HR. Prevalence of Dietary Supplement Use by Athletes: Systematic Review and Meta-Analysis. Sports Med. 2016;46: 103–123. doi:10.1007/s40279-015-0387-7

36. Manner T, Katz DP, Askanazi J. The antinociceptive effects of branched-chain amino acids: Evidence for their ability to potentiate morphine analgesia. Pharmacol Biochem Be. 1996;53: 449–454. doi:10.1016/0091-3057(95)02016-0

37. Wagenmakers A. Amino Acid Metabolism, Muscular Fatigue and Muscle Wasting. Int J Sports Med. 1992;13: S110–S113. doi:10.1055/s-2007-1024611

38. Aquilani R, Boselli M, Boschi F, Viglio S, Iadarola P, Dossena M, et al. Branched-Chain Amino Acids May Improve Recovery From a Vegetative or Minimally Conscious State in Patients With Traumatic Brain Injury: A Pilot Study. Arch Phys Med Rehab. 2008;89: 1642–1647. doi:10.1016/j.apmr.2008.02.023

39. Aquilani R, Iadarola P, Contardi A, Boselli M, Verri M, Pastoris O, et al. Branched-Chain Amino Acids Enhance the Cognitive Recovery of Patients With Severe Traumatic Brain Injury. Arch Phys Med Rehab. 2005;86: 1729–1735. doi:10.1016/j.apmr.2005.03.022

40. Blomstrand E. Amino acids and cenral fatigue. Amino Acids. 2001;20: 25–34.

41. Blomstrand E, Hassmen P, Ekblom B, Newsholme EA. Administration of branched-chain amino acids during sustained exercise--effects on performance and on plasma concentration of some amino acids. European journal of applied physiology. 1991;63: 83–88.

42. Hasseman P, Blomstrand E, Ekblom B, Newsholme EA. Branched-chain amino acid supplementation during 30-km competitive run: mood and cognitive performance. Nutrition. 1994;10: 405–410.

43. Cerra FB, Mazuski J, Teasley K, Nuwer N, Lysne J, Shronts E, et al. Nitrogen retention in critically ill patients is proportional to the branched chain amino acid load. Crit Care Med. 1983;11: 775–778. doi:10.1097/00003246-198310000-00003

44. Fernstrom JD. Large neutral amino acids: dietary effects on brain neurochemistry and function. Amino Acids. 2013;45: 419–430. doi:10.1007/s00726-012-1330-y

45. Horst D, Grance ND, Conn HO, Schiff E, Schenker S, Viteri A, et al. Comparison of dietary protein with an oral, branched chain-enriched amino acid supplement in chronic portal-systemic encephalopathy: a randomized controlled trial. Hepatology. 1984;4: 279–287.

46. James JH. Branched chain amino acids in heptatic encephalopathy. Am J Surg. 2002;183: 424–429. doi:10.1016/s0002-9610(02)00808-5

47. Marchesini G, Bianchi G, Merli M, Amodio P, Panella C, Loguercio C, et al. Nutritional supplementation with branched-chain amino acids in advanced cirrhosis: a double-blind, randomized trial. Gastroenterology. 2003;124: 1792–1801. doi:10.1016/s0016-5085(03)00323-8

48. Mori M, Adachi Y, Mori N, Kurihara S, Kashiwaya Y, Kusumi M, et al. Double-blind crossover study of branched-chain amino acid therapy in patients with spinocerebellar degeneration. J Neurol Sci. 2002;195: 149–152. doi:10.1016/s0022-510x(02)00009-6

49. Ozgultekın A, Turan G, Durmus Y, Dıncer E, Akgun N. Comparison of the efficacy of parenteral glutamine and branched-chain amino acid solutions given as extra supplements in parallel to the enteral nutrition in head trauma. E-spen European E-journal Clin Nutrition Metabolism. 2008;3: e211–e216. doi:10.1016/j.eclnm.2008.05.006

50. Plauth M, Egberts E-H, Hamster W, Török M, Müller PH, Brand O, et al. Long-term treatment of latent portosystemic encephalopathy with branched-chain amino acids A double-blind placebo-controlled crossover study. J Hepatol. 1993;17: 308–314. doi:10.1016/s0168-8278(05)80210-7

51. Richardson MA, Bevans ML, Weber JB, Gonzalez JJ, Flynn CJ, Amira L, et al. Branched chain amino acids decrease tardive dyskinesia symptoms. Psychopharmacology. 1999;143: 358–364. doi:10.1007/s002130050959

52. Scarna A, Gijsman H, McTavish SF, Harmer CJ, Cowen PJ, Goodwin GM. Effects of a branched-chain amino acid drink in mania. British Journal of Psychiatry. 2003;182: 210–213.

53. Skeie B, Kvetan V, Gil KM, Rothkopf MM, Newsholme EA, Askanazi J. Branch-chain amino acids. Crit Care Med. 1990;18: 549–571. doi:10.1097/00003246-199005000-00019

54. Tandan R, Bromberg MB, Forshew D, Fries TJ, Badger GJ, Carpenter J, et al. A controlled trial of amino acid therapy in amyotrophic lateral sclerosis. Neurology. 1996;47: 1220–1226. doi:10.1212/wnl.47.5.1220

55. Elliott JE, Keil AT, Mithani S, Gill JM, O’Neil ME, Cohen AS, et al. Dietary Supplementation With Branched Chain Amino Acids to Improve Sleep in Veterans With Traumatic Brain Injury: A Randomized Double-Blind Placebo-Controlled Pilot and Feasibility Trial. Frontiers Syst Neurosci. 2022;16: 854874. doi:10.3389/fnsys.2022.854874

56. Corwin DJ, Myers SR, Arbogast KB, Lim MM, Elliott JE, Metzger KB, et al. Head Injury Treatment With Healthy and Advanced Dietary Supplements: A Pilot Randomized Controlled Trial of the Tolerability, Safety, and Efficacy of Branched Chain Amino Acids in the Treatment of Concussion in Adolescents and Young Adults. J Neurotrauma. 2024. doi:10.1089/neu.2023.0433

57. Morin CM, Belleville G, Bélanger L, Ivers H. The Insomnia Severity Index: psychometric indicators to detect insomnia cases and evaluate treatment response. Sleep. 2011;34: 601–608.

58. Johns MW. A new method for measuring daytime sleepiness: the Epworth sleepiness scale. Sleep. 1991;14: 540–545.

59. Chasens ER, Ratcliffe SJ, Weaver TE. Development of the FOSQ-10: A Short Version of the Functional Outcomes of Sleep Questionnaire. Sleep. 2009;32: 915–919. doi:10.1093/sleep/32.7.915

60. Mastin DF, Bryson J, Corwyn R. Assessment of sleep hygiene using the Sleep Hygiene Index. J Behav Med. 2006;29: 223–227. doi:10.1007/s10865-006-9047-6

61. Hays RD, Bjorner JB, Revicki DA, Spritzer KL, Cella D. Development of physical and mental health summary scores from the patient-reported outcomes measurement information system (PROMIS) global items. Qual Life Res. 2009;18: 873–880. doi:10.1007/s11136-009-9496-9

62. Cella D, Yount S, Rothrock N, Gershon R, Cook K, Reeve B, et al. The Patient-Reported Outcomes Measurement Information System (PROMIS). Med Care. 2007;45: S3–S11. doi:10.1097/01.mlr.0000258615.42478.55

63. Vanderploeg RD, Cooper DB, Belanger HG, Donnell AJ, Kennedy JE, Hopewell CA, et al. Screening for Postdeployment Conditions. J Head Trauma Rehab. 2014;29: 1–10. doi:10.1097/htr.0b013e318281966e

64. Blevins CA, Weathers FW, Davis MT, Witte TK, Domino JL. The Posttraumatic Stress Disorder Checklist for *DSM-5* (PCL-5): Development and Initial Psychometric Evaluation. Journal of Traumatic Stress. 2015;28: 489–498. doi:10.1002/jts.22059

65. Kroenke K, Spitzer RL, Williams JB. The PHQ-9: validity of a brief depression severity measure. J Gen Intern Med. 2001;16: 606–613.

66. Ustün TB, Chatterji S, Kostanjsek N, Rehm J, Kennedy C, Epping-Jordan J, et al. Developing the World Health Organization Disability Assessment Schedule 2.0. B World Health Organ. 2010;88: 815–823. doi:10.2471/blt.09.067231

